# Several forms of SARS-CoV-2 RNA can be detected in wastewaters : implication for wastewater-based epidemiology and risk assessment

**DOI:** 10.1101/2020.12.19.20248508

**Authors:** S. Wurtzer, P. Waldman, A. Ferrier-Rembert, G. Frenois-Veyrat, JM. Mouchel, M. Boni, Y. Maday, OBEPINE consortium, V. Marechal, L. Moulin

## Abstract

The ongoing global pandemic of coronavirus disease 2019 (COVID-19) caused by severe acute respiratory syndrome coronavirus 2 (SARS-CoV-2) has been a public health emergency of international concern. Although SARS-CoV-2 is considered to be mainly transmitted by inhalation of contaminated droplets and aerosols, SARS-CoV-2 is also detected in human feces and in raw wastewaters suggesting that other routes of infection may exist. Monitoring SARS-CoV-2 genomes in wastewaters has been proposed as a complementary approach for tracing the dynamics of virus transmission within human population connected to wastewater network. The understanding on SARS-CoV-2 transmission through wastewater surveillance, the development of epidemic modeling and the evaluation of SARS-CoV-2 transmission from contaminated wastewater are largely limited by our knowledge on viral RNA genome persistence and virus infectivity preservation in such an environment. Using an integrity based RT-qPCR assay this study led to the discovery that SARS-CoV-2 RNA can persist under several forms in wastewaters, which provides important information on the presence of SARS-CoV-2 in raw wastewaters and associated risk assessment.

**Graphical Abstract:** 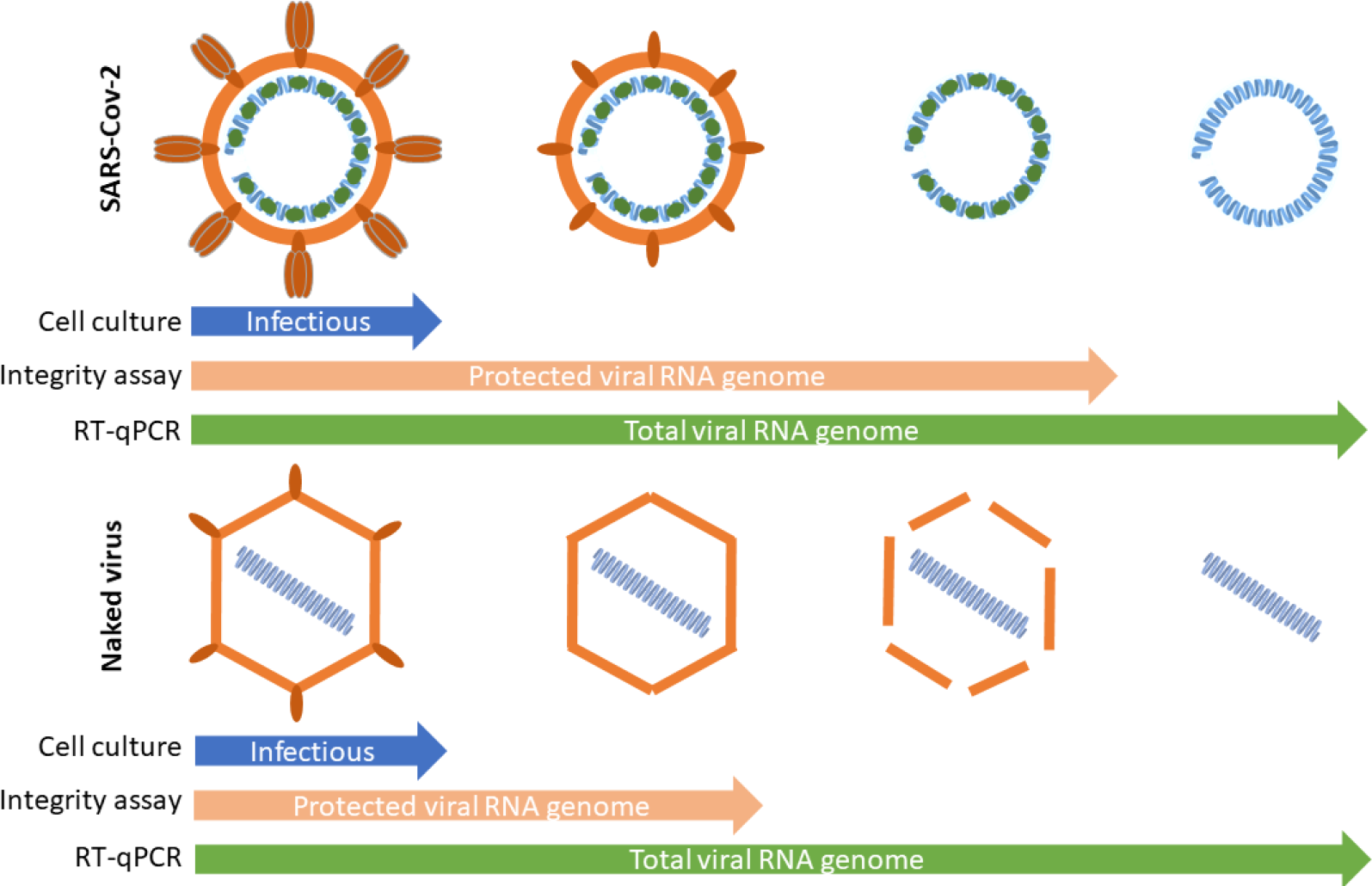

## Introduction

Coronaviruses (CoVs) belong to coronaviridae, a large family of enveloped single-stranded positive RNA viruses. CoVs are usually considered as moderate pathogens for humans. Four of them (229E, NL63, OC43, HKU1) are responsible for seasonnal common cold or mild respiratory infections. However, three novel and highly pathogenic CoVs recently emerged in human population causing severe zoonotic diseases i.e. Severe Acute Respiratory Syndrome (SARS)(Peiris et al., 2003), Middle East Respiratory Syndrome (MERS)(Zaki et al., 2012) and more recently COronaVIrus Disease-19 (COVID-19). SARS-CoV-2, the etiological agent of COVID-19(Huang et al., 2020; Zhou et al., 2020; Zhu et al., 2020), is responsible for a pandemia that caused at least 67 million cases and more than 1.5 million deaths so far (John Hopkins university data by december 7^th^, 2020). Although SARS-CoV-2 transmission mainly occurs by direct transmission through inhalation of contaminated respiratory droplets or through contaminated aerosols or surfaces(WHO, n.d.), the potential for alternative transmission pathway should not be underestimated. Indeed, large amounts of viral RNA have been identified in human stools from infected patients presenting with severe COVID-19 symptoms which occasionaly led to the isolation of infectious virus from feces(Chen et al., 2020; Holshue et al., 2020; Huang et al., 2020; Lescure et al., 2020; Pan et al., 2020a; Tang et al., 2020; Wang et al., 2020; Wölfel et al., 2020; Wu et al., 2020; Xiao et al., 2020; Zhang et al., 2020). SARS-CoV-2 can also be detected in stools from asymptomatic carriers with a largely unknown prevalence(Tang et al., 2020). This likely reflects SARS-CoV-2 replication in the gut(Luz et al., 2020). Accordingly high level of viral RNA have been detected in wastewaters in different countries and potential cases of transmission via wastewater have been reported(Yeo et al., 2020a; Yuan et al., 2020). In addition to the risk of exposure for sewage workers, wastewaters containing potentially infectious SARS-CoV-2 may enter the aquatic environment via wastewater discharge thus potentially resulting in pollution of surface waters (Kumar et al., 2020; Naddeo and Liu, 2020; Rimoldi et al., 2020; Wurtzer et al., 2020a)and to a lesser extent groundwaters. Such a pollution could locally affect the quality of water ressources used for the production of water intended to human consumption. Moreover, the persistence of infectious virus in treated effluents of wastewater treatment plant could cause problems for agricultural activities through the reuse of treated wastewater or the spreading of sludge(Balboa et al., 2020). Consequently, the contamination of wastewater by SARS-CoV-2 raises the same concerns as human seasonal enteric viruses(Okoh et al., 2010).

The monitoring of SARS-CoV-2 genomes in raw wastewater was successfully used for estimating the dynamics of viral pandemic in population linked to a wastewater network(Medema et al., 2020; Nemudryi, n.d.; Randazzo et al., 2020; Wurtzer et al., 2020b). However many questions remain to be answered to better assess the risk of transmission of SARS-CoV-2 through wastewaters(Elsamadony et al., 2021; Lodder and de Roda Husman, 2020). Indeed RT-qPCR protocoles that are currently used can not distinguish between partial or full-length, virion associated or free viral genomes(Prevost et al., 2016). It is commonly admitted that enveloped viruses are less persistent in hydric matrices and less resistant to inactivation treatments than naked viruses(WHO, n.d.). Gundy and collaborators showed that human seasonal coronavirus survival in tap water and wastewater was strongly reduced compared to poliovirus. The survival ranged from days to weeks depending on the surrogate virus, type of water and temperature(Casanova et al., 2009; Casanova and Weaver, 2015; Gundy et al., 2009). An experimental study showed that SARS-CoV-1 stability under an infectious form was only 2 days at 20 °C, but 14 days at 4 °C(Wang et al., 2005). So far only a few studies investigated SARS-CoV-2 stability on solid surfaces(Chin et al., 2020; van Doremalen et al., 2020) or in water matrix(Bivins et al., 2020). If the decay of SARS-CoV-2 infectivity appears to be different according to the nature of matrix, these few studies agreed on the sensitivity to heat. Moreover they suggested that SARS-CoV-2 could be more persistent than other coronaviruses (seasonal and epidemic CoV) and more resistant to harsh condition(Aboubakr et al., 2020; van Doremalen et al., 2020). Conversely risk assessment for SARS-CoV-2 was mainly based on results obtained for other coronaviruses or for SARS-CoV-2 surrogates (enteric viruses or bacteriophage indicators)(Rosa et al., 2020; Silverman and Boehm, 2020; Ye et al., 2016). So far, despite the presence of SARS-CoV-2 RNA in raw wastewaters, no infectious virus was isolated from the same samples, suggesting that the detection of viral RNA overestimated the risk of infection(Rimoldi et al., 2020).

The present work intended to evaluate SARS-CoV-2 stability both under an infectious form or by quantifying viral RNA in wastewaters. We first demonstrated that SARS-CoV-2 RNA can be quantified without significant loss in wastewaters samples for up to 7 days at 4°C or 20°C, suggesting that viral RNA is largely protected from environnemental degradation. This led us to combine cell culture isolation and integrity based RT-qPCR assay to investigate the status of viral RNA in wastewater samples(Prevost et al., 2016). We propose that SARS-CoV-2 genomes can exist under three different states at least: genomic RNA protected within an infectious particle, genomic RNA protected in a non-infectious structure, free total or partial genomic RNA. SARS-CoV-2 persistence and integrity were compared to an enteric virus – Coxsackievirus B5 – that is commonly found in feces and wastewater. The analysis of 87 raw wastewater samples collected from April to July 2020 in Paris area confirmed that total viral RNA can be detected under both a protected and an unprotected form.

## Material and methods

### Virus stock preparation

Coxsackievirus B5 (CV-B5) was cultivated on confluent monolayer cultures of Buffalo Green Monkey kidney (BGMK) cells at 37°C with 5% CO_2_. Cells were grown in Dulbecco’s Modified Eagle’s Medium (DMEM) high glucose (Dutscher) supplemented with 2% fetal bovine serum (PanBiotech), non-essential amino acids (Dutscher), penicillin (100 U/ml) and streptomycin (100 µg/ml) (PanBiotech). The supernatant was clarified by centrifugation at 2,000 *x g* for 15 min, then ultracentrifuged at 150,000 *x g* at 4 °C for 2 hours through a 40 % sucrose cushion. The pellet was resuspended in 1x phosphate-buffered saline (PBS) pH 7.4. Further purification was performed by ultracentrifugation on cesium chloride gradient at 100,000 *x g* for 18 hours. The fraction containing the viruses was desalted with Vivaspin 20 (Sartorius) concentrators according the manufacturer’s recommandations. Viruses were stored at - 80 °C before using.

SARS-CoV-2, strain SARS Cov-2 20/0001 (BetaCoV/France/IDF0372/2020/SARS-CoV-2 isolated by Pasteur Institute, France), was cultivated on confluent monolayer cultures of VERO cells, kindly provided by Dr. Le Gouil and Pr. Vabret (Virology laboratory of universitary hospital of Caen, France) at 37°C with 5% CO_2_. Cells were grown in Dulbecco’s Modified Eagle’s Medium GlutaMAX (Gibco) supplemented with penicillin (50 U/mL) and streptomycin (50µg/mL), TPCK trypisin (1µg/mL) without fetal bovin serum. The supernatant, collected after cytopathic effect observation, was clarified by centrifugation at 2,000 *x g* for 15 min and stored at - 80 °C before using.

### Detection of SARS-CoV-2 in raw wastewater

Raw wastewater samples were homogenized, then 11 ml were centrifugated at 200 000 × g for 1 hour at +4°C using a XPN80 Coulter Beckman ultracentrifuge equipped with a swing rotor (SW41Ti). Viral pellets were resuspended in 200 μL of PBS 1X buffer as previsouly described by Wurtzer & al.

### Spiking assays

Five raw wastewater samples were collected in july 2020 in different WWTP and scored negative for SARS-CoV-2 and enterovirus genome. These <24h old samples were centrifugated at 4,000 xg for 15 min for removing the largest particles and supernatants were filtred on membrane with 0,45µm porosity. The filtrates were stored at +4°C and used within the following 24h.

CV-B5 or SARS-CoV-2 were spiked in the filtrated samples. Virus titration was immediately done after spiking or after incubation at 4°C or 20°C for the indicated period of time. As a control, spiking experiments were done in DMEM. Virus infectivity, virus integrity and viral RNA detection were assessed after incubation by endpoint dilution assay, PMAxx-RT-qPCR and RT-qPCR respectively.

### Virus quantification by endpoint dilution assay

Infectious viruses (CV-B5 and SARS-CoV-2) were titrated by standard 10-fold dilutions in 96-well plates on VERO E6 cells (ATCC® CRL-1586™) (10^5^ cells per well), with twelve replicates per dilution. After a 6-day incubation, cytopathic effects were observed and positive wells were counted. Viral titer was estimated using the Spearman-Kårber method. The results are expressed as 50% tissue culture infective dose (TCID_50_) per ml.

### Virus integrity Assay

Each sample was mixed with Propidium monoazide (PMAxx), an intercalating dyes that binds only to free accessible sites within nucleic acids and after photoactivation, making them unable to be amplified by RT-qPCR. Briefly PMAxx was added at 100 µM final concentration. The samples were incubated on ice in the dark for 30 min and then photoactivated at using PhastBlue system (GenIUL, Spain) for 15 min. Samples were extracted as follow.

### Viral RNA detection

#### Spiking assays

The spiked samples were lysed by adding two volumes of TRIZOL (Lifetechnologies) and extracted using QIAsymphony DSP/ Pathogen kit on a QIAsymphony automated extractor (QIAGEN) according to a modified manufacturer’s protocol for handling larger volumes.

#### Environmental samples

The viral concentrate was lysed and extracted using Qiasymphony PowerFecal Pro kit on a QIAsymphony automated extractor (QIAGEN) according to a modified manufacturer’s protocol. Extracted nucleic acids were filtered through OneStep PCR inhibitor removal kit (Zymoresearch) according the manufacturer’s instructions for handling larger volumes.

#### Viral RNA titration

The RT-qPCR primers and PCR conditions used herein have been previously described(Corman et al., 2020). The amplification was done using Fast virus 1-step Master mix 4x (Lifetechnologies). Detection and quantification were carried on the gene E by RT-qPCR. Positive results were confirmed by amplification of viral RNA-dependent RNA polymerase (RdRp) and nucleoprotein genes. An internal positive control (IPC) was added to evaluate the presence of residual inhibitors. The IPC consists in a plasmid containing beta-acting gene flanked by enterovirus-specific primers(Wurtzer et al., 2014). The detection limit was estimated to be around 10 genome units per amplification reaction.

The quantification was performed using a standard curve based on full-length amplicon cloned into pCR2.1 plasmid (Invitrogen, #452640). Amplification reaction and fluorescence detection were performed on Viia7 Real Time PCR system (Lifetechnologies).

### Statistical analysis and plots

All statistical analysis and plots were done using GraphPad Prism 9.0 software. For comparison based on spiked samples (figure 2), the quantifications were compared between the different conditions using one-way ANOVA and Tukey’s multiple comparisons test. Comparisons between total vRNA and protected RNA (figure 4A) were performed using Wilcoxson matched-pair test and comparisons of ratio pRNA/vRNA (figure 4B) were tested using Kruskal-Wallis test and Dunn’s multiple comparisons test.

## Results

### Stability of total viral RNA (vRNA) in wastewater samples

The quantification of SARS-CoV-2 genome in wastewater has been proposed as an alternative strategy to monitor the dynamics of pandemic SARS-CoV-2 virus. However, this approach is highly dependent on the persistence of SARS-CoV-2 RNA in wastewaters. In addition, it is of upmost importance to provide convenient tools to distinguish free viral RNA and virion-associated RNA as a first approach to evaluate the concentration of infectious virus particle in matrix from which SARS-CoV-2 is technically difficult to isolate, such as stools or wastewaters. Since viral genomes are protected by viral proteins and surrounded by a cell-derived enveloped in infectious particles, we assumed that we could distinguish between free and protected viral genomes using an integrity RT-qPCR based assay.

Briefly, two 1L-raw wastewater samples were collected by the 3^rd^ (sample S1) and the 7^th^ (sample S2) of april 2020 in Greater Paris area, a period when SARS-CoV-2 genomes were easily detected(Wurtzer et al., 2020b). Samples were analyzed less than 24h after the time of the sampling (day 0). The rest of each sample was split into 2 parts and stored at +4°C or +20°C for 10 days and 12 days respectively. Total SARS-Cov-2 viral RNA (vRNA) and protected viral RNA (pRNA) were quantified by RT-qPCR. As shown on figure 1, less than 10 % of the total viral RNA was under a protected form. SARS-CoV-2 vRNA and pRNA concentrations were relatively stable for 7 (S1) and 12 (S2) days respectively at +4°C while they were slighly less stable when stored at +20°C.

**Figure 1.**
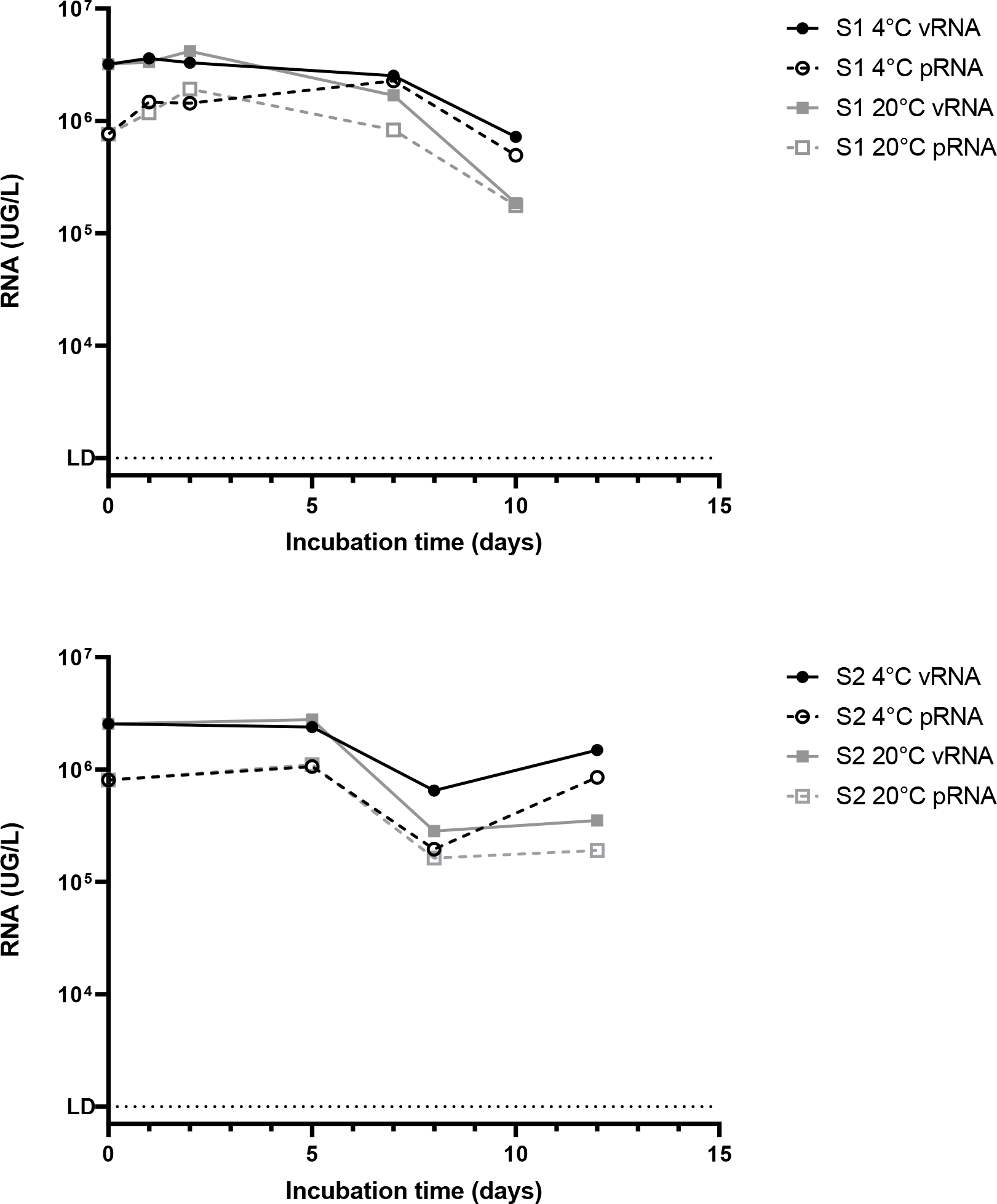
Persistence of total or protected SARS-CoV-2 RNA of in two raw wastewater samples. Two naturally SARS-CoV-2 contaminated wastewater samples (S1 and S2) were independently incubated at +4°C or +20°C for several days. Total viral RNA (vRNA, filled forms) and protected viral RNA (pRNA, open forms) were quantified by RT-qPCR and by an integrity-based RT-PCR respectively.

**Figure 2.**
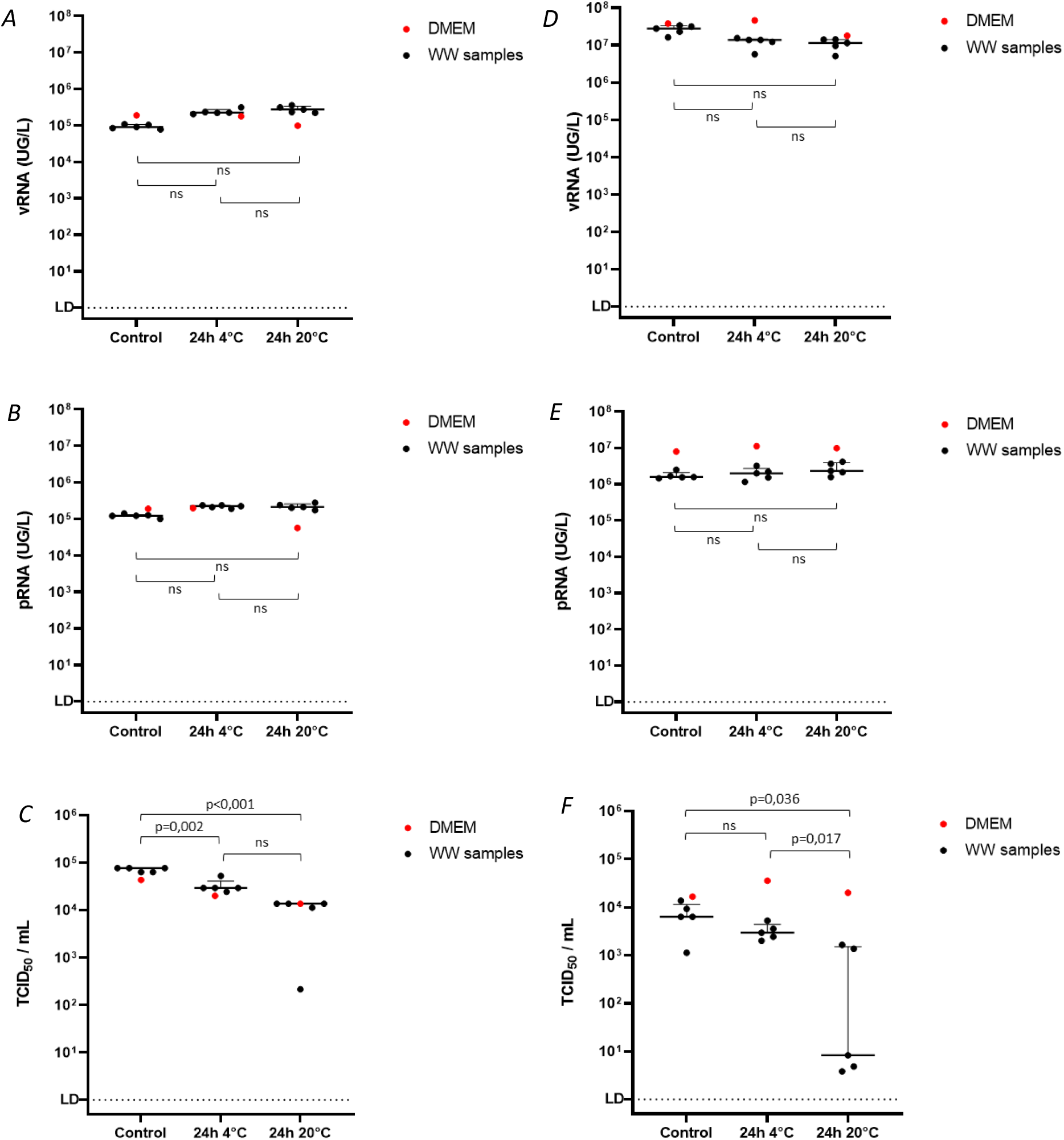
Stability of total viral RNA, protected viral RNA and infectious SARS-CoV-2 and coxsackievirus B5 in spiked wastewater samples. Five wastewater samples were spiked with infectious virus and incubated for 24h at +4°C or +20°C. DMEM was used as a control of matrix. Total viral RNA (vRNA) of CV-B5 (panel A) or SARS-CoV-2 (panel D) were quantified by RT-qPCR. Protected RNA (pRNA) of CV-B5 (panel B) or SARS-CoV-2 (panel E) were quantified using an integrity-based RT-PCR, as described. Infectious particles (TCID50) of CV-B5 (panel C) or SARS-CoV-2 (panel F) were titrated by cell culture.

### Comparing coxsackievirus B5 and SARS-CoV-2 persistence in raw wastewater

Infectious enteric virus such as coxsackievirus B5 are commonly found in wastewaters, but the ability of enveloped virus, like SARS-CoV-2, to persist under an infectious form is still debated. To address this question the persistence of SARS-CoV-2 in raw wastewater was compared to that of coxsackievirus B5 (CV-B5) using three different indicators namely the quantification of total RNA (vRNA), protected viral RNA (pRNA) and infectious particles (TCID_50_). Five raw wastewater samples, that were negative for SARS-CoV-2 and enterovirus genome by RT-qPCR (data not shown) were used. The detection of infectious virus, vRNA and pRNA was performed after spiking each sample with infectious SARS-CoV-2 or CV-B5.

CV-B5 vRNA and pRNA were quantified at similar concentrations in raw wastewaters (WW) or in cell culture medium (DMEM) when analysis was done immediately after spiking (control) or after 24h-incubation à +4°C or +20°C (figure 2A, 2B). This result was expected since CV-B5 particles were purified to homogeneity on sucrose gradient, which efficiently separates encapsidated RNA from free RNA. Infectivity of CV-B5 was not significantly altered after 24h-incubation at +4°C, while it only sightly decreased (<1-log) after a 24-incubation at +20°C (figure 2C). One WW samples dramatically affected the virus infectivity (>2-log). Strikingly, pRNA was only 10% of total vRNA for SARS-CoV-2 suggesting that unpurified SARS-CoV-2 preparation contains only a minor part of intact particles. This result was further confirmed by the relatively low level of infectivity of the viral stock (figure 2C). As before pRNA was highly stable whereas total SARS-CoV-2 total vRNA partly decreased over time in all conditions (figure 2D and 2E). As importantly SARS-CoV-2 infectivity was strongly (>3-log) or moderately reduced in 3 out of 5 WW samples and 2 over 5 samples respectively. The decrease in TCID_50_ was about 1-log in all samples after a 24h-incubation at +4°C. Since no similar observation was made on samples containing DMEM, this suggested that SARS-CoV-2 infectivity is strongly reduced in wastewaters likely depending of their chemical and/or microbial composition (figure 2F).

### Temperature-based inactivation unrevealed different status for viral RNA

Temperature is known to affect viruses in the environment albeit to very different extent(Bertrand et al., 2012). Heat inactivation is commonly used for studying virus survival in water. In low temperature range (<50°C), the inactivation of naked viruses mainly comes from the denaturation of capsids (Waldman et al., 2020, 2017). However little is known concerning enveloped viruses. Therefore, we intended to evaluate more precisely the effect of temperature on SARS-CoV-2 using CV-B5 as a control. For this purpose, we first exposed samples spiked with infectious SARS-CoV-2 and CV-B5 to increasing temperature for 10 minutes. Then we evaluated the effect of the treatment on infectious particles or total RNA stability (vRNA). Viral genome protection was evaluated as before by an integrity RT-qPCR based assay (pRNA).

CV-B5 infectiosity was preserved up to 42°C and then dramatically decreased up to 70°C, as previously described by Waldman and co-authors(Waldman et al., 2017). pRNA and vRNA were stable up to 50 and 70°C respectively in culture medium, although RNA integrity significantly decreased at a lower temperature in wastewater (figure 3 A).

**Figure 3.**
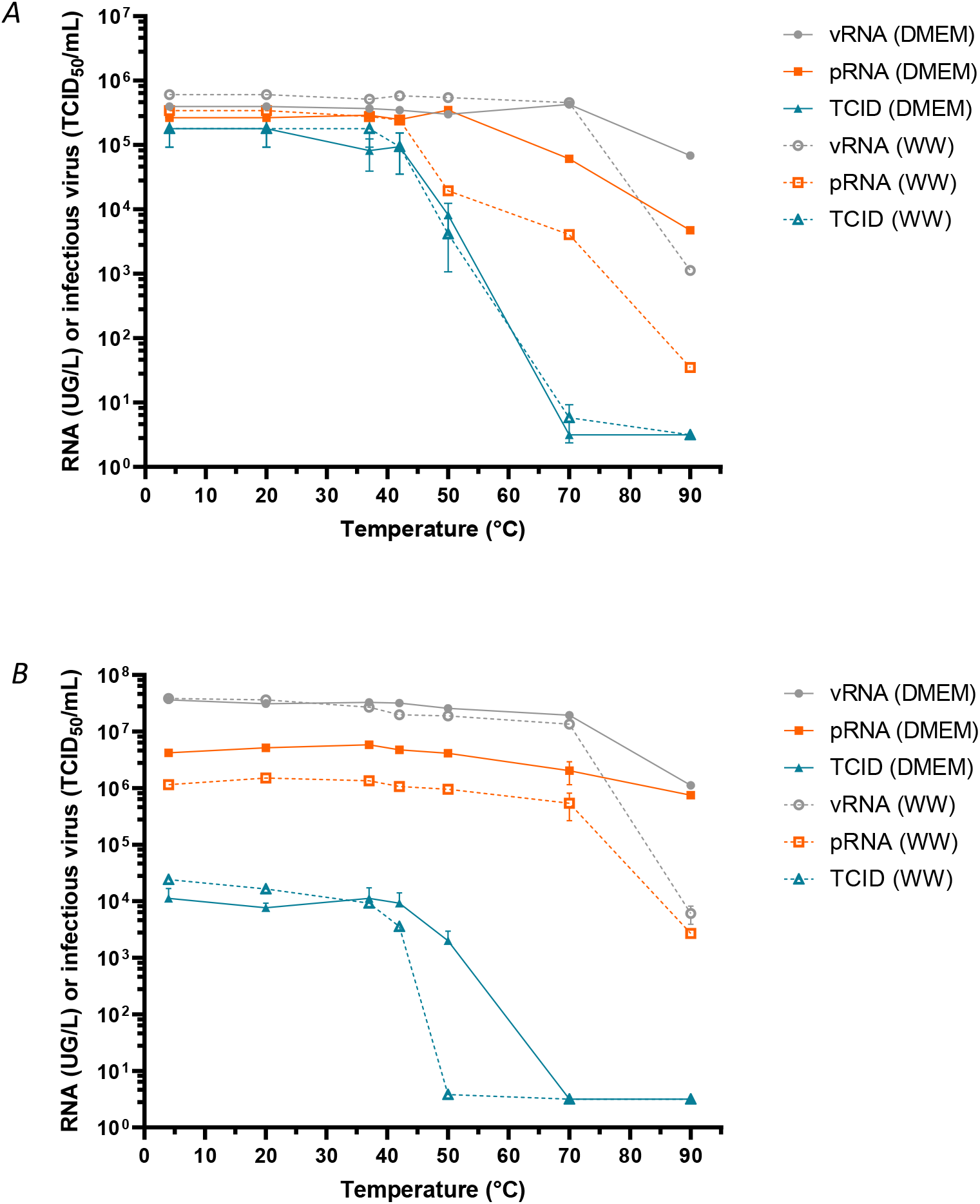
Stability to heat of total viral RNA, protected viral RNA and infectious SARS-CoV-2 and coxsackievirus B5 in spiked wastewater. Wastewater samples were spiked with infectious CV-B5 particles (panel A) or infectious SARS-CoV-2 particles (panel B) and incubated for 10 min at various temperatures. Total viral RNA (vRNA) was quantified by RT-qPCR, protected RNA (pRNA) was quantified by an integrity-based RT-PCR, as described, and infectious virus (TCID50) was titrated by cell culture.

**Figure 4.**
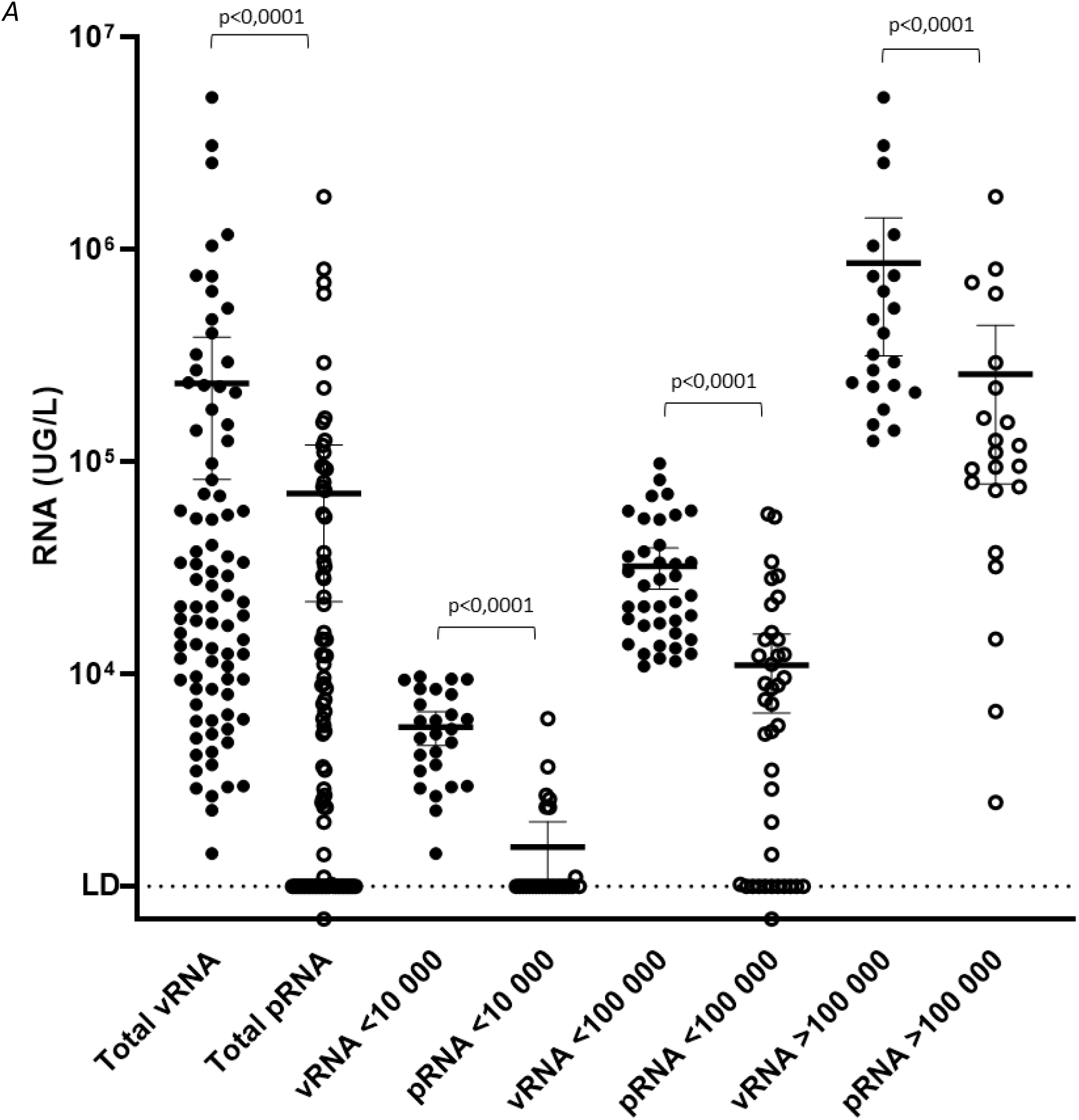
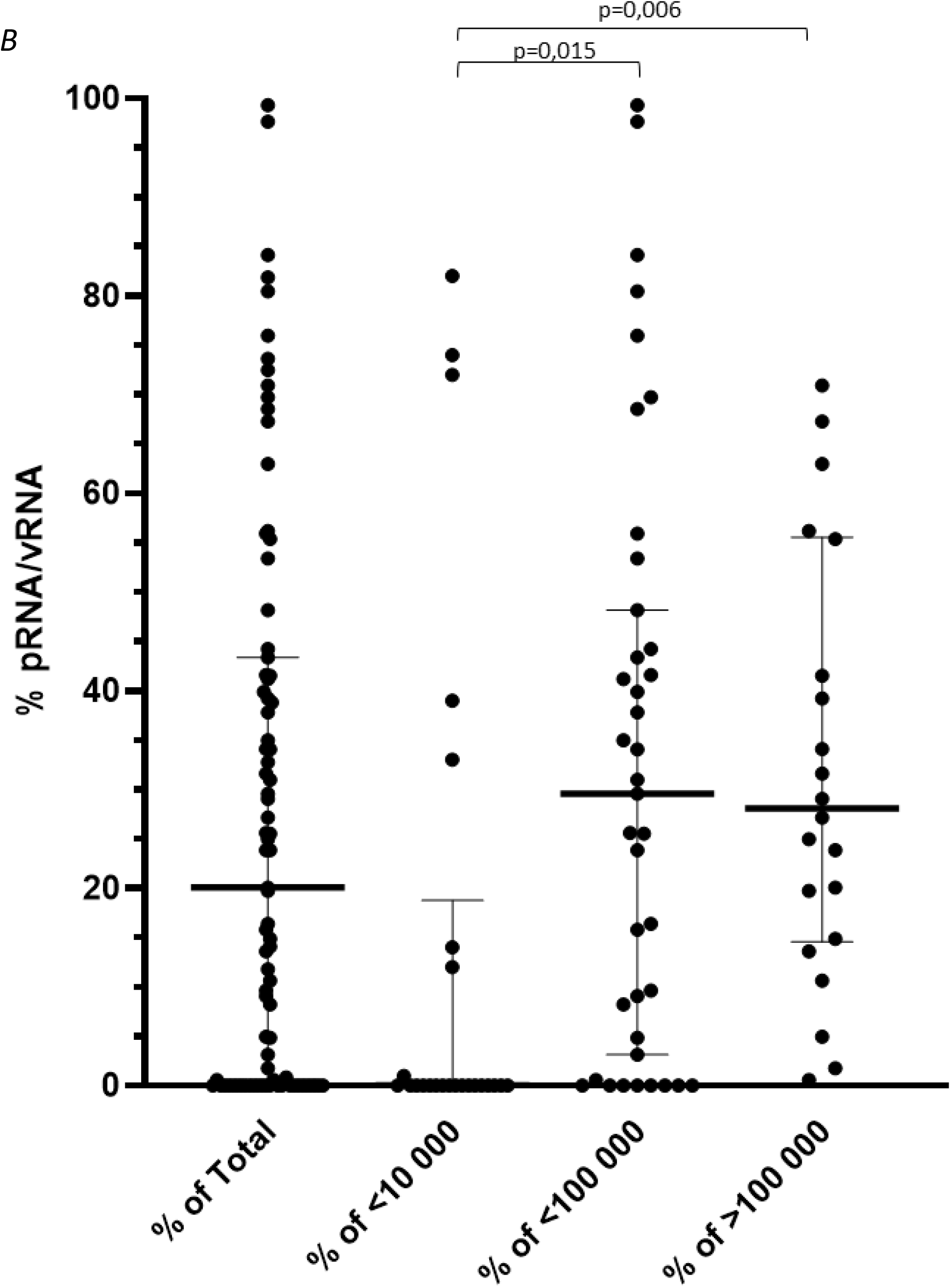
Relative proportion of protected vs unprotected SARS-CoV-2 genomes in raw wastewaters collected in Greater Paris area. Raw wastewater samples (n=87) from four WWTP were analyzed for SARS-CoV-2 genome by RT-qPCR (vRNA, filled circle) and using integrity assay (pRNA, open circle). The concentration (UG/L) was plotted on the panel A, the median values and interquartiles (25-75%) are indicated. The pRNA/vRNA ratio indicating the percentage of protected RNA over total viral RNA, is plotted for each sample on panel B. The median values and interquartiles (25-75%) are indicated.

In the same conditions, SARS-CoV-2 viability was not significantly affected until 42°C. A marked reduction in infectiosity was observed both in wastewaters and culture medium that was not related with a decrease in vRNA nor pRNA (figure 3 B). In both culture medium and wastewater samples, reduction of vRNA paralleled pRNA reduction although reduction in vRNA and pRNA was stronger in wastewater sample.

Altogether, these experiments indicated that SARS-CoV-2 viral genomes could exist under three different forms at least: protected within infectious particles, protected within non-infectious particles or in a ribonucleoprotein complex and as free/unprotected viral RNA.

### Estimating the relative proportion of protected vs unprotected SARS-CoV-2 genomes in raw wastewater

Total and protected viral RNA were quantified in 87 raw wastewater samples that were collected from April to July 2020 in Greater Paris area. vRNA and pRNA concentrations ranged from 1.4×10^3^ to 5.2×10^6^ GU/L and from 0.7×10^3^ to 1.8×10^6^ genome units/L respectively (figure 4A). Total viral RNA were significantly higer than protected RNA in each sample (p<0.0001). The pRNA/vRNA ratio was comprised between 0 and 100%, with a median value of 20.1%. In wastewater samples with vRNA concentrations <100,000 (n=39) and >100,000 GU/L (n=22), the median ratio was 29,6% (max=99,3%) and 28.1% (max=100%) respectively. This ratio was significantly lower in samples with low genome concentration (n=26; <10,000 GU/L; median ratio = 0%; max = 18.8%) compared to <100,000 GU/L and >100,000 GU/L samples (p=0.015 and p=0.006 respectively) (figure 4B).

## Discussion

Wastewater-based epidemiology has been widely used over the world for monitoring the spreading of SARS-CoV-2(Medema et al., 2020; Randazzo et al., 2020; Wurtzer et al., 2020b) in human populations as well as other waterborn viruses such as poliovirus (WHO, 2003) and other enteric viruses(Prevost et al., 2015). A large panel of methods has been developed with various performances. SARS-CoV-2 is detected in feces of about 50% of infected people, mainly with no or moderate symptoms(Lescure et al., 2020; Pan et al., 2020b; Tang et al., 2020). It has been proposed that the presence of SARS-CoV-2 genomes in raw wastewater could reflect the virus excreted by infected people, whether they are symptomatic or not. It is of upmost importance to confirm this assumption in order to propose mathematical models that could correlate viral load in wastewaters with other individual epidemiological parameters. Modeling viral dynamics greatly depends on the quality of the analysis, but also on the half-life of total viral RNA in raw wastewater. In this study, we showed that total viral RNA (vRNA) concentration in raw wastewater was stable for at least 7 days provided that the samples were stored at +4°C until analysis, which is in agreement with previous work(Bivins et al., 2020). Importantly, freezing water samples had a negative impact on the relevance of the measurment (data not shown), at least in our protocol. Such a delay is important to be taken into consideration to organize campains from the sampling to the analysis, including transportation to specialized laboratories. Although our study was performed on a limited number of samples, the results suggested that vRNA concentration was not dramatically affected by the composition of wastewater samples over 24h-incubation time, a period of time that is compatible with the travel of the viral genomes from emission of human faeces to raw wastewater sampling at the inlet of WWTP. As importantly SARS-CoV-2 vRNA detection was unaffected in a range of temperature comprised between +4°C and at least 40°C. Altogether these results suggested that the measurement of SARS-CoV-2 vRNA concentration in wastewater is a relevant indicator of the effective level of the viral genomes excreted by infected people that is only moedartely affected by temperature and travel time.

The detection of SARS-CoV-2 genomes in stools and subsequently in wastewaters raises several other important concerns concerning the risk of transmission. RT-qPCR assays have been designed to detect specific regions of the viral genomes whatever the quantified RNA is extracted from infectious particle or not. Therefore, these approaches provide an obvious overestimate of the effective concentration of infectious viral particles within stools and wastewaters. Even though sewage is an unsanitary environment for many reasons, sewers and operators of wastewater treatment plant worried about the occupational risk of infection by SARS-CoV-2. As recently underlined by WHO(WHO, n.d.), SARS-CoV-2 is a respiratory virus whose main routes of transmission are respiratory (inhalation of contaminated dropplets) and contact (with contaminated surfaces). However, if SARS-CoV-2 infection via contaminated wastewater was not unambiguously demonstrated, this possibility cannot be ruled out(Yeo et al., 2020b). To that respect, let us note that genetic evidences and case clustering led Yuan and coworkers to suggest that sewage may be a possible transmission vehicle for SARS-CoV-2(Gormley, 2020; Kang et al., 2020; Yuan et al., 2020). Enveloped viruses were commonly thought to be less resistant than naked virus. Due to the possible presence of detergent and other chemical agents that may degrade viral envelop, raw wastewater might be highly detrimental to the persistence of infectious SARS-Cov2 particles. Trials have been done unsuccessfully in order to isolate and cultivate SARS-CoV-2 from fresh wastewater samples(Rimoldi et al., 2020), meaning that SARS-CoV-2 might be simply non-infectious, or that cell culture system was not adapted for such highly chemically or microbiologically contaminated samples(Cashdollar and Wymer, 2013). Efforts for concentrating and isolating infectious viruses from hydric environment are usually successful for naked virus that are less sensitive to chemicals. In this study, infectious SARS-CoV-2 was spiked in negative wastewater samples and viable viruses were quantified up to 24 hours, without pretreatment of sample before cultivation.

Whereas such an exposure only midly affected coxsackievirus B5 viability, SARS-CoV-2 infectivity was clearly affected at 20°C depending on the nature of the sample. These results are in agreement with previous work(Bivins et al., 2020). We brought here additional evidences that sample temperature had a strong impact on virus viability since the SARS-CoV-2 infectivity was not significantly modified at +4°C for 24h whereas it is slightly affected at 20°C. Both viruses infectivity was fully preserved up to 42°C for shorter incubation times (10 min). In all conditions, infectious virus persisted up to 24 hours at least in wastewater samples. Previous studies reported that infectious SARS-CoV-2 could persist for up to 28 days on various supports (glass, plastic or stainless steel for example) (Riddell et al., 2020; van Doremalen et al., 2020). An effect of temperature on viral infectivity was already reported when virus was adsorbed on solid surfaces(Riddell et al., 2020) or in transportation medium(Chin et al., 2020). More recently Bivins and collaborators brought first elements to evaluate SARS-CoV-2 viability in wastewaters and provided evidences that SARS-CoV-2 viral RNA persisted for longer period of time than infectious particles(Bivins et al., 2020).

The present study confirmed that evaluating total vRNA widely overestimated the number of infectious particles within wastewaters. Nevertheless, the relatively long persistence of SARS-CoV-2 genomes was surprising with regards to its supposed fragility compared to surrogates. Whether the regions that are amplified by RT-qPCR came from total or partial genomes cannot be assessed by such assays. A tool for assessing the integrity of naked virus particles already showed that genome of naked RNA viruses can be protected from degradation by the capsid, a structure that remains non-permeable to intercalating dye. Our comparative study on SARS-CoV-2 and CV-B5 demonstrated that viral genomes can be found in multiple states i.e. infectious protected, non-infectious protected and unprotected forms. Unpublished data showed that such dyes (Ethidium monoazide or propidium monoazide) targeted secondary structures within single stranded RNA (hairpins or IRES for picornaviruses)(Wurtzer et al., 2018). In addition previous study showed that capsid integrity is lost at 42°C for CV-B5, with a maximum access of SyBR green II to viral RNA at 50°C(Waldman et al., 2017). In the case of coronaviruses, a lipid layer protects the RNA genome that is closely associated to nucleoproteins. The lipid layer is probably very labile in wastewater, which may contain detergent residues for example, and unstable at high temperature. Nonetheless integrity measurments showed that vRNA remained protected from intercalating dye up to 70°C-incubation. These results suggested other structures such as viral nucleoproteins may limit access of the dye to SARS-CoV-2 RNA, in addition to the viral envelop. It is to note that SARS-CoV-2 and CV-B5 shared a similar profil of sensitivity to temperature, although SARS-CoV-2 genome appeared to be better protected than CV-B5 genomes. This assay was used on a large panel of samples, confirming that less than 30% of the viral genomes on the average were under a protected form in wastewater samples. Considering that infectious particles correspond only to a subfraction of protected genomes, as illustrated by spiking experiments, it can be considered that risk assement for viral infection through wastewaters should be better evaluated using an integrity based assay if systematic cell culture isolation cannot be done. Such a technique could also be used to evaluate the relative fraction of protected genomes in other matrix such as in sputums or stools of infected patients or for other enveloped viruses, such as influenza virus(Chan et al., 2009; Hirose, 2016).

## Contribution

SW and PW made the virus measurements; FRA, FVG and MB participed and facilitated the infectivity assay in BSL3 laboratory; YM and JMM facilitated wastewater sampling; SW, PW, VM and LM for the redaction of the manuscript; YM, JMM and MB for critical discussion.

Obepine consortium includes Isabelle Bertrand, Soizick Le Guyarder, Christophe Gantzer, Mickael Boni, Vincent Maréchal, Yvon Maday, Jean-Marie Mouchel, Laurent Moulin and Sébastien Wurtzer.

## Data Availability

our study did not use any human clinical data, but all data are available if they need to be transfered.

## References

Aboubakr, H.A., Sharafeldin, T.A., Goyal, S.M., 2020. Stability of SARS-CoV-2 and other coronaviruses in the environment and on common touch surfaces and the influence of climatic conditions: A review. Transbound Emerg Dis tbed.13707. https://doi.org/10.1111/tbed.13707

Balboa, S., Mauricio-Iglesias, M., Rodríguez, S., Martínez-Lamas, L., Vasallo, F.J., Regueiro, B., Lema, J.M., 2020. The fate of SARS-CoV-2 in wastewater treatment plants points out the sludge line as a suitable spot for incidence monitoring (preprint). Epidemiology. https://doi.org/10.1101/2020.05.25.20112706

Bertrand, I., Schijven, J.F., Sánchez, G., Wyn-Jones, P., Ottoson, J., Morin, T., Muscillo, M., Verani, M., Nasser, A., de Roda Husman, A.M., Myrmel, M., Sellwood, J., Cook, N., Gantzer, C., 2012. The impact of temperature on the inactivation of enteric viruses in food and water: a review. J. Appl. Microbiol. 112, 1059–1074. https://doi.org/10.1111/j.1365-2672.2012.05267.x

Bivins, A., Greaves, J., Fischer, R., Yinda, K.C., Ahmed, W., Kitajima, M., Munster, V.J., Bibby, K., 2020. Persistence of SARS-CoV-2 in Water and Wastewater. Environ. Sci. Technol. Lett. 7, 937–942. https://doi.org/10.1021/acs.estlett.0c00730

Casanova, L., Rutala, W.A., Weber, D.J., Sobsey, M.D., 2009. Survival of surrogate coronaviruses in water. water research 6.

Casanova, L.M., Weaver, S.R., 2015. Inactivation of an Enveloped Surrogate Virus in Human Sewage. Environmental Science 3.

Cashdollar, J.L., Wymer, L., 2013. Methods for primary concentration of viruses from water samples: a review and meta-analysis of recent studies. J Appl Microbiol 115, 1–11. https://doi.org/10.1111/jam.12143

Chan, M.C.W., Lee, N., Chan, P.K.S., Leung, T.F., Sung, J.J.Y., 2009. Fecal detection of influenza A virus in patients with concurrent respiratory and gastrointestinal symptoms. Journal of Clinical Virology 45, 208–211. https://doi.org/10.1016/j.jcv.2009.06.011

Chen, C., Gao, G., Xu, Y., Pu, L., Wang, Q., Wang, Liming, Wang, W., Song, Y., Chen, M., Wang, Linghang, Yu, F., Yang, S., Tang, Y., Zhao, L., Wang, H., Wang, Y., Zeng, H., Zhang, F., 2020. SARS-CoV-2–Positive Sputum and Feces After Conversion of Pharyngeal Samples in Patients With COVID-19. Ann Intern Med. https://doi.org/10.7326/M20-0991

Chin, A.W.H., Chu, J.T.S., Perera, M.R.A., Hui, K.P.Y., Yen, H.-L., Chan, M.C.W., Peiris, M., Poon, L.L.M., 2020. Stability of SARS-CoV-2 in different environmental conditions. The Lancet Microbe 1, e10. https://doi.org/10.1016/S2666-5247(20)30003-3

Corman, V.M., Landt, O., Kaiser, M., Molenkamp, R., Meijer, A., Chu, D.K.W., Bleicker, T., Brunink, S., Schneider, J., Schmidt, M.L., Mulders, D.G.J.C., Haagmans, B.L., van der Veer, B., van den Brink, S., Wijsman, L., Goderski, G., Romette, J.-L., Ellis, J., Zambon, M., Peiris, M., Goossens, H., Reusken, C., Koopmans, M.P.G., Drosten, C., 2020. Detection of 2019 novel coronavirus (2019-nCoV) by real-time RT-PCR. Euro Surveill 25. https://doi.org/10.2807/1560-7917.ES.2020.25.3.2000045

Elsamadony, M., Fujii, M., Miura, T., Watanabe, T., 2021. Possible transmission of viruses from contaminated human feces and sewage: Implications for SARS-CoV-2. Science of The Total Environment 755, 142575. https://doi.org/10.1016/j.scitotenv.2020.142575

Gormley, M., 2020. SARS-CoV-2: The Growing Case for Potential Transmission in a Building via Wastewater Plumbing Systems. Ann Intern Med 173, 1020–1021. https://doi.org/10.7326/M20-6134

Gundy, P.M., Gerba, C.P., Pepper, I.L., 2009. Survival of Coronaviruses in Water and Wastewater. Food Environ Virol 1, 10. https://doi.org/10.1007/s12560-008-9001-6

Hirose, R., 2016. Long-term detection of seasonal influenza RNA in faeces and intestine. Clinical Microbiology and Infection 7.

Holshue, M.L., DeBolt, C., Lindquist, S., Lofy, K.H., Wiesman, J., Bruce, H., Spitters, C., Ericson, K., Wilkerson, S., Tural, A., Diaz, G., Cohn, A., Fox, L., Patel, A., Gerber, S.I., Kim, L., Tong, S., Lu, X., Lindstrom, S., Pallansch, M.A., Weldon, W.C., Biggs, H.M., Uyeki, T.M., Pillai, S.K., 2020. First Case of 2019 Novel Coronavirus in the United States. The New England Journal of Medicine 8.

Huang, C., Wang, Y., Li, X., Ren, L., Zhao, J., Hu, Y., Zhang, L., Fan, G., Xu, J., Gu, X., Cheng, Z., Yu, T., Xia, J., Wei, Y., Wu, W., Xie, X., Yin, W., Li, H., Liu, M., Xiao, Y., Gao, H., Guo, L., Xie, J., Wang, G., Jiang, R., Gao, Z., Jin, Q., Wang, J., Cao, B., 2020. Clinical features of patients infected with 2019 novel coronavirus in Wuhan, China. The Lancet 395, 497–506. https://doi.org/10.1016/S0140-6736(20)30183-5

Kang, M., Wei, J., Yuan, J., Guo, J., Zhang, Y., Hang, J., Qu, Y., Qian, H., Zhuang, Y., Chen, X., Peng, X., Shi, T., Wang, J., Wu, J., Song, T., He, J., Li, Y., Zhong, N., 2020. Probable Evidence of Fecal Aerosol Transmission of SARS-CoV-2 in a High-Rise Building. Ann Intern Med 173, 974–980. https://doi.org/10.7326/M20-0928

Kumar, M., Thakur, A.K., Mazumder, P., Kuroda, K., Mohapatra, S., Rinklebe, J., Ramanathan, Al., Cetecioglu, Z., Jain, S., Tyagi, V.K., Gikas, P., Chakraborty, S., Tahmidul Islam, M., Ahmad, A., Shah, A.V., Patel, A.K., Watanabe, T., Vithanage, M., Bibby, K., Kitajima, M., Bhattacharya, P., 2020. Frontier review on the propensity and repercussion of SARS-CoV-2 migration to aquatic environment. Journal of Hazardous Materials Letters 1, 100001. https://doi.org/10.1016/j.hazl.2020.100001

Lescure, F.-X., Bouadma, L., Nguyen, D., Parisey, M., Wicky, P.-H., Behillil, S., Gaymard, A., Bouscambert-Duchamp, M., Donati, F., Le Hingrat, Q., Enouf, V., Houhou-Fidouh, N., Valette, M., Mailles, A., Lucet, J.-C., Mentre, F., Duval, X., Descamps, D., Malvy, D., Timsit, J.-F., Lina, B., van-der-Werf, S., Yazdanpanah, Y., 2020. Clinical and virological data of the first cases of COVID-19 in Europe: a case series. The Lancet Infectious Diseases 20, 697–706. https://doi.org/10.1016/S1473-3099(20)30200-0

Lodder, W., de Roda Husman, A.M., 2020. SARS-CoV-2 in wastewater: potential health risk, but also data source. Lancet Gastroenterol Hepatol. https://doi.org/10.1016/S2468-1253(20)30087-X

Luz, B.B. da, Oliveira, N.M.T. de, Santos, I.W.F. dos, Paza, L.Z., Braga, L.L.V. de M., Platner, F. da S., Werner, M.F. de P., Fernandes, E.S., Maria-Ferreira, D., 2020. An overview of the gut side of the SARS-CoV-2 infection. Intest Res. https://doi.org/10.5217/ir.2020.00087

Medema, G., Heijnen, L., Elsinga, G., Italiaander, R., Brouwer, A., 2020. Presence of SARS-Coronavirus-2 RNA in Sewage and Correlation with Reported COVID-19 Prevalence in the Early Stage of the Epidemic in The Netherlands. Environmental Science 6.

Naddeo, V., Liu, H., 2020. Editorial Perspectives: 2019 novel coronavirus (SARS-CoV-2): what is its fate in urban water cycle and how can the water research community respond? Environ. Sci.: Water Res. Technol. 6, 1213–1216. https://doi.org/10.1039/D0EW90015J

Nemudryi, A., n.d. Temporal Detection and Phylogenetic Assessment of SARS-CoV-2 in Municipal Wastewater. OPEN ACCESS 11.

Okoh, A.I., Sibanda, T., Gusha, S.S., 2010. Inadequately Treated Wastewater as a Source of Human Enteric Viruses in the Environment. Int. J. Environ. Res. Public Health 19.

Pan, Y., Zhang, D., Yang, P., Poon, L.L.M., Wang, Q., 2020a. Viral load of SARS-CoV-2 in clinical samples. The Lancet Infectious Diseases 20, 411–412. https://doi.org/10.1016/S1473-3099(20)30113-4

Pan, Y., Zhang, D., Yang, P., Poon, L.L.M., Wang, Q., 2020b. Viral load of SARS-CoV-2 in clinical samples. The Lancet Infectious Diseases 20, 411–412. https://doi.org/10.1016/S1473-3099(20)30113-4

Peiris, J.S.M., Lai, S.T., Poon, L.L.M., Guan, Y., Yam, L.Y.C., Lim, W., Nicholls, J., Yee, W.K.S., Yan, W.W., Cheung, M.T., Cheng, V.C.C., Chan, K.H., Tsang, D.N.C., Yung, R.W.H., Ng, T.K., Yuen, K.Y., 2003. Coronavirus as a possible cause of severe acute respiratory. THE LANCET 361, 8.

Prevost, B., Goulet, M., Lucas, F.S., Joyeux, M., Moulin, L., Wurtzer, S., 2016. Viral persistence in surface and drinking water: Suitability of PCR pre-treatment with intercalating dyes. Water Res 91, 68–76. https://doi.org/10.1016/j.watres.2015.12.049

Prevost, B., Lucas, F.S., Goncalves, A., Richard, F., Moulin, L., Wurtzer, S., 2015. Large scale survey of enteric viruses in river and waste water underlines the health status of the local population. Environ Int 79, 42–50. https://doi.org/10.1016/j.envint.2015.03.004

Randazzo, W., Cuevas-Ferrando, E., Sanjuan, R., Domingo-Calap, P., Sanchez, G., 2020. Metropolitan wastewater analysis for COVID-19 epidemiological surveillance. International Journal of Hygiene and Environmental Health 5.

Riddell, S., Goldie, S., Hill, A., Eagles, D., Drew, T.W., 2020. The effect of temperature on persistence of SARS-CoV-2 on common surfaces. Virol J 17, 145. https://doi.org/10.1186/s12985-020-01418-7

Rimoldi, S.G., Stefani, F., Gigantiello, A., Polesello, S., Comandatore, F., Mileto, D., Maresca, M., Longobardi, C., Mancon, A., Romeri, F., Pagani, C., Cappelli, F., Roscioli, C., Moja, L., Gismondo, M.R., Salerno, F., 2020. Presence and infectivity of SARS-CoV-2 virus in wastewaters and rivers. Science of The Total Environment 744, 140911. https://doi.org/10.1016/j.scitotenv.2020.140911

Rosa, G.L., Bonadonna, L., Lucentini, L., Kenmoe, S., Suffredini, E., 2020. Coronavirus in water environments: Occurrence, persistence and concentration methods - A scoping review. Water Research 12.

Silverman, A.I., Boehm, A.B., 2020. Systematic Review and Meta-Analysis of the Persistence and Disinfection of Human Coronaviruses and Their Viral Surrogates in Water and Wastewater. Environ. Sci. Technol. Lett. 7, 544–553. https://doi.org/10.1021/acs.estlett.0c00313

Tang, B., Wang, X., Li, Q., Bragazzi, N.L., Tang, S., Xiao, Y., Wu, J., 2020. Estimation of the Transmission Risk of the 2019-nCoV and Its Implication for Public Health Interventions. Journal of Clinical Medicine 9, 462. https://doi.org/10.3390/jcm9020462

van Doremalen, N., Bushmaker, T., Morris, D.H., Holbrook, M.G., Gamble, A., Williamson, B.N., Tamin, A., Harcourt, J.L., Thornburg, N.J., Gerber, S.I., Lloyd-Smith, J.O., de Wit, E., Munster, V.J., 2020. Aerosol and Surface Stability of SARS-CoV-2 as Compared with SARS-CoV-1. N Engl J Med 382, 1564–1567. https://doi.org/10.1056/NEJMc2004973

Waldman, P., Lucas, F.S., Varrault, G., Moulin, L., Wurtzer, S., 2020. Hydrophobic Organic Matter Promotes Coxsackievirus B5 Stabilization and Protection from Heat. Food Environ Virol. https://doi.org/10.1007/s12560-019-09418-9

Waldman, P., Meseguer, A., Lucas, F., Moulin, L., Wurtzer, S., 2017. Interaction of Human Enteric Viruses with Microbial Compounds: Implication for Virus Persistence and Disinfection Treatments. Environ. Sci. Technol. 51, 13633–13640. https://doi.org/10.1021/acs.est.7b03875

Wang, W., Xu, Y., Gao, R., Lu, R., Han, K., Wu, G., Tan, W., 2020. Detection of SARS-CoV-2 in Different Types of Clinical Specimens. JAMA. https://doi.org/10.1001/jama.2020.3786

Wang, X.-W., Li, J.-S., Jin, M., Zhen, B., Kong, Q.-X., Song, N., Xiao, W.-J., Yin, J., Wei, W., Wang, G.-J., Si, B., Guo, B.-Z., Liu, C., Ou, G.-R., Wang, M.-N., Fang, T.-Y., Chao, F.-H., Li, J.-W., 2005. Study on the resistance of severe acute respiratory syndrome-associated coronavirus. Journal of Virological Methods 7.

WHO, 2003. Guidelines for environmental surveillance of poliovirus circulation. (No.= WHO/V&B/03.03).

WHO, n.d. Transmission of SARS-CoV-2: implications for infection prevention precautions (No. WHO/2019-nCoV/Sci_Brief/Transmission_modes/2020.3).

WHO, n.d. Water, sanitation, hygiene, and waste management for the COVID-19 virus (No. WHO/2019-nCoV/IPC_WASH/2020.3).

Wölfel, R., Corman, V.M., Guggemos, W., Seilmaier, M., Zange, S., Müller, M.A., Niemeyer, D., Jones, T.C., Vollmar, P., Rothe, C., Hoelscher, M., Bleicker, T., Brünink, S., Schneider, J., Ehmann, R., Zwirglmaier, K., Drosten, C., Wendtner, C., 2020. Virological assessment of hospitalized patients with COVID-2019. Nature 581, 465–469. https://doi.org/10.1038/s41586-020-2196-x

Wu, Y., Guo, C., Tang, L., Hong, Z., Zhou, J., Dong, X., Yin, H., Xiao, Q., Tang, Y., Qu, X., Kuang, L., Fang, X., Mishra, N., Lu, J., Shan, H., Jiang, G., Huang, X., 2020. Prolonged presence of SARS-CoV-2 viral RNA in faecal samples. Lancet Gastroenterol Hepatol. https://doi.org/10.1016/S2468-1253(20)30083-2

Wurtzer, S., Marechal, V., Mouchel, J., Maday, Y., Teyssou, R., Richard, E., Almayrac, J., Moulin, L., 2020a. Time course quantitative detection of SARS-CoV-2 in Parisian wastewaters correlates with COVID-19 confirmed cases. (preprint). Epidemiology. https://doi.org/10.1101/2020.04.12.20062679

Wurtzer, S., Marechal, V., Mouchel, J., Maday, Y., Teyssou, R., Richard, E., Almayrac, J., Moulin, L., 2020b. Evaluation of lockdown impact on SARS-CoV-2 dynamics through viral genome quantification in Paris wastewaters (preprint). Epidemiology. https://doi.org/10.1101/2020.04.12.20062679

Wurtzer, S., Prevost, B., Lucas, F.S., Moulin, L., 2014. Detection of enterovirus in environmental waters: a new optimized method compared to commercial real-time RT-qPCR kits. J Virol Methods 209, 47–54. https://doi.org/10.1016/j.jviromet.2014.08.016

Wurtzer, S., Waldman, P., Moulin, L., 2018. New insights for optimizing molecular detection of infectious viruses.

Xiao, F., Sun, J., Xu, Y., Li, F., Huang, X., Li, H., Zhao, Jingxian, Huang, J., Zhao, Jincun, 2020. Infectious SARS-CoV-2 in Feces of Patient with Severe COVID-19. Emerg. Infect. Dis. 26, 1920–1922. https://doi.org/10.3201/eid2608.200681

Ye, Y., Ellenberg, R.M., Graham, K.E., Wigginton, K.R., 2016. Survivability, Partitioning, and Recovery of Enveloped Viruses in Untreated Municipal Wastewater. Environ. Sci. Technol. 50, 5077–5085. https://doi.org/10.1021/acs.est.6b00876

Yeo, C., Kaushal, S., Yeo, D., 2020a. Enteric involvement of coronaviruses: is faecal–oral transmission of SARS-CoV-2 possible? The Lancet Gastroenterology & Hepatology 5, 335–337. https://doi.org/10.1016/S2468-1253(20)30048-0

Yeo, C., Kaushal, S., Yeo, D., 2020b. Enteric involvement of coronaviruses: is faecal–oral transmission of SARS-CoV-2 possible? The Lancet Gastroenterology & Hepatology 5, 335–337. https://doi.org/10.1016/S2468-1253(20)30048-0

Yuan, J., Chen, Z., Gong, C., Liu, H., Li, B., Li, K., Chen, X., Xu, C., Jing, Q., Liu, G., Qin, P., Liu, Y., Zhong, Y., Huang, L., Zhu, B.-P., Yang, Z., 2020. Sewage as a Possible Transmission Vehicle During a Coronavirus Disease 2019 Outbreak in a Densely populated Community: Guangzhou, China, April 2020. Clinical Infectious Diseases ciaa 1494. https://doi.org/10.1093/cid/ciaa1494

Zaki, A.M., van Boheemen, S., Bestebroer, T.M., Osterhaus, A.D.M.E., Fouchier, R.A.M., 2012. Isolation of a novel coronavirus from a man with pneumonia in Saudi Arabia. N Engl J Med 367, 1814–1820. https://doi.org/10.1056/NEJMoa1211721

Zhang, Y., Chen, C., Zhu, S., Shu, C., Wang, D., Song, J., Song, Y., Zhen, W., Feng, Z., Wu, G., 2020. Isolation of 2019-nCoV from a stool specimen of a laboratory-confirmed case of the coronavirus disease 2019 (COVID-19). China CDC Weekly 2, 123–124.

Zhou, P., Yang, X.-L., Wang, X.-G., Hu, B., Zhang, L., Zhang, W., Si, H.-R., Zhu, Y., Li, B., Huang, C.-L., Chen, H.-D., Chen, J., Luo, Y., Guo, H., Jiang, R.-D., Liu, M.-Q., Chen, Y., Shen, X.-R., Wang, X., Zheng, X.-S., Zhao, K., Chen, Q.-J., Deng, F., Liu, L.-L., Yan, B., Zhan, F.-X., Wang, Y.-Y., Xiao, G.-F., Shi, Z.-L., 2020. Discovery of a novel coronavirus associated with the recent pneumonia outbreak in humans and its potential bat origin (preprint). Microbiology. https://doi.org/10.1101/2020.01.22.914952

Zhu, N., Zhang, D., Wang, W., Li, X., Yang, B., Song, J., Zhao, X., Huang, B., Shi, W., Lu, R., Niu, P., Zhan, F., Ma, X., Wang, D., Xu, W., Wu, G., Gao, G.F., Tan, W., 2020. A Novel Coronavirus from Patients with Pneumonia in China, 2019. N Engl J Med 382, 727–733. https://doi.org/10.1056/NEJMoa2001017

